# UK-based mental health professionals’ perceptions of genetic and environmental risk for psychiatric conditions

**DOI:** 10.1101/2025.09.24.25336580

**Authors:** Helena L. Davies, Jessica Mundy, Karla Mohoric, Molly R. Davies, Thalia C. Eley, Gerome Breen, Evangelos Vassos

**Affiliations:** Social Genetic & Developmental Psychiatry Centre, Institute of Psychiatry Psychology & Neuroscience, King’s College London, UK; Department of Health Services Research & Policy, Faculty of Public Health, London School of Hygiene & Tropical Medicine, London, UK; National Institute for Health and Care Research (NIHR) Maudsley Biomedical Research Centre, South London and Maudsley NHS Foundation Trust, London, UK

**Author notes:** Corresponding author;, Social Genetic and Developmental Psychiatry Centre, Memory Lane, London, SE5 8AF. Joint first authors (Helena L. Davies, Jessica Mundy).

## Abstract

**Objective:** Psychiatric conditions arise from a complex interplay of genetics and environment. Many people struggle to interpret genetic risk, leading to distress and inadequately informed decisions. Mental health professionals are often the first point of contact for patients with questions about psychiatric genetic risk. We aimed to uncover UK-based psychiatric professionals’ knowledge and perceptions of psychiatric genetic risk, comparing those with and without a medical degree to identify training gaps.

**Methods:** Healthcare professionals (n=152) were recruited via email and social media. An online survey assessed: 1) genetic knowledge using the 6-item International Genetic Literacy and Attitudes Survey (iGLAS), 2) confidence discussing genetic and environmental risk, 3) perceived contributions of genetic and environmental factors to psychiatric conditions, and 4) how often patients and relatives ask questions about genetic or environmental risk. We compared those with a medical degree (n=56) to those without (n=96) via linear regressions.

**Results:** One-quarter indicated patients always or often asked about genetic risk for their psychiatric condition. Whilst participants’ genetic knowledge was good (mean iGLAS score=4.36 out of 6), 10% believed at least one psychiatric condition was caused by only environmental or only genetic factors. Only 30% feel confident discussing genetic risk with patients and their relatives. Participants with a medical degree demonstrated significantly greater genetic knowledge (mean iGLAS score=5.02, SD=0.91) than those without (mean=3.97, SD=1.31; β=1.05, 95% CI=0.64, 1.47, *p*<0.001), and were more confident discussing genetic risk (β=0.60, 95% CI=0.20, 1.01, *p*=0.004), however this latter finding was non-significant after controlling for sex. Confidence discussing environmental risk was not linked to training background.

**Conclusions:** Patients and their relatives are curious about genetic risk, yet professionals’ confidence discussing this was low, especially among those without medical degrees. We highlight a specific training gap related to genetics and support calls for accessible psychiatric genetics education for mental health professionals.

## Introduction

Psychiatric conditions are complex and polygenic; social and environmental factors combine and interact with thousands - possibly tens of thousands - of rare and common genetic variants in a person’s genome which each contribute a tiny amount to a person’s overall risk^1^. Individual genetic risk information is becoming widely available to the public and clinicians, most often in the form of polygenic scores, which represents a person’s genetic risk for a trait or disease, based on common DNA variants included in the score. These common variants have been identified through genome-wide association studies, a method that looks for genomic markers associated with a particular trait or disease. For some somatic diseases, such as breast cancer^2^ and rheumatoid arthritis^3^, polygenic scores are being extensively evaluated for their clinical utility to identify at-risk groups and prevention (e.g., by combining polygenic scores with information on other known risk factors in screening programmes). However, polygenic scores explain just a small proportion of the variance in psychiatric risk, only capture risk conferred by common genetic variants, and do not include structural, repeat, or rare genetic variation. For example, the median area under the receiver operating characteristic curve for the schizophrenia polygenic score - one of the most powerful in psychiatry - for predicting schizophrenia diagnosis is only 0.72^4^. Therefore, polygenic scores are not currently used clinically for predicting the development of a psychiatric condition. However, as psychiatric genetics research advances and the cost of whole genome sequencing decreases, the combination of polygenic scores, other genetic variation, as well as other clinical risk information, could aid clinical decision-making in psychiatry in the future^5–8^.

### Example cases of genetic testing for psychiatric conditions

Nonetheless, in 2023, the US Food and Drug Administration approved a genetic test of opioid use disorder for adolescents being considered for opioid pain medications to treat acute pain, for example following surgery^9^, and private companies have already begun offering customers information on their psychiatric genetic risk via direct-to-consumer (DTC) genetic testing kits. In June 2024, 23andMe (a company now owned by Regeneron after declaring bankruptcy) added bipolar disorder to its list of psychiatric conditions that it provides genetic risk information for, joining depression, panic attacks, and anxiety, despite the very low predictive accuracy of the information available. Additionally, some private fertility companies based in the US now offer polygenic embryo screening for complex psychiatric and cognitive traits such as schizophrenia^10^ and IQ^11^, which has been criticised, as outlined in a 2022 statement from the International Society of Psychiatric Genetics (ISPG)^12^.

### (Lack of) public understanding of psychiatric genetics

As genetic data become increasingly commercialised and part of public consciousness, it is crucial that individuals understand the complex and non-deterministic nature of genetic risk for psychiatric conditions. Psychiatric genetic risk is not easily communicated, especially in simple electronic formats such as those offered by DTC genetic testing companies. Genetic risk is not well understood by the public^13^. Only one-quarter of DTC genetic test users who accessed a third party site to generate polygenic scores demonstrated a complete understanding and interpretation of these scores; concerningly, those with a lower understanding displayed significantly greater negative emotions related to the genomic test results^14^. Such negative consequences of misunderstanding genetic risk, like concerns about having children, have been found previously^15–20^.

### The role of mental health professionals

Genetic counselling - a process through which a trained professional helps an individual to better understand and adapt to the medical, psychological, and familial implications of genetic contributions to disease - is the ideal course of action^21^. However, as of 2019, there were only 310 genetic counsellors in the UK and most work in somatic diseases^22^. Thus, individuals who are confused or concerned about, e.g., their family history, their children’s risk of developing a psychiatric condition, or results from a DTC genetic test, may turn to their primary healthcare practitioner or other health professionals for reassurance or clarification^23^. In fact, 10% of 960 US-based child and adolescent psychiatrists reported having been brought a polygenic score by a patient or their family member^24^. A review by the ISPG Residency Education Committee outlined that the rise in DTC genetic testing means that psychiatrists need to understand genetics in order to accurately and responsibly address potential concerns about results, as well as knowing when to consult genetic counsellors and medical geneticists^25^. However, awareness, knowledge of, and training in genetics among mental health professionals is low^26–29^; even amongst genetic counsellors, confidence in providing meaningful psychiatric genetic counselling is similarly low^30^. Self-rated competencies of discussing psychiatric genetic risk differ significantly depending on training background, with medical geneticists rating themselves significantly more competent than genetic counsellors or psychiatrists^31^.

As our understanding of psychiatric genetics advances and the integration of genetics into clinical care becomes increasingly possible^6,32–34^, alongside the sharp rise in DTC genetic testing companies, the need for mental health professionals to have accurate and responsible conversations with patients about the heritable nature of psychiatric conditions is more pertinent now than ever. With that in mind, we investigated the level of general genetic knowledge among UK-based mental health professionals and the extent to which they understand and feel confident discussing genetic and environmental risk for psychiatric conditions with their patients and their patients’ relatives. Second, we investigated mental health professionals’ views on psychiatric genetic counselling. Finally, because medical training emphasises the biological aspects of disease in preparation for prescribing, we compared the level of knowledge and confidence in professionals with and without a medical degree, to identify potential gaps in training programmes.

## Methods

### Recruitment

Participants were invited to complete an anonymous online survey between 19th March 2022 and 3rd February 2023. We distributed the survey via email to 62 academic and clinical organisations and mental health charities across the UK and advertised it on social media sites including Instagram and Twitter (now X). Interested participants were directed to an online information sheet and consent form before completing the online survey via Qualtrics. Upon completion, all participants were eligible to enter a prize draw to win an iPad.

### Survey

The survey took 10-15 minutes to complete. Questions were designed by the PerPsych study team with input from clinical psychologists, psychiatrists, and genetic counsellors. A copy of the full survey can be found in the Supplementary Material. The survey began with questions about the participants’ demographic characteristics (e.g., age, sex, gender, race) and professional background including job role, time since qualifying, whether they worked privately or for the National Health Service (NHS) or both, their current professional setting, and which psychiatric or neurodevelopmental conditions they currently worked with.

We assessed participants’ knowledge of genetics with the shortened version of the International Genetic Literacy and Attitudes Survey (iGLAS^35^; 6-item version obtained via personal communication, see Supplementary Material) as well as their confidence in discussing genetic and environmental risk with patients/clients. Next, we asked participants their perception of the genetic and environmental contributions to each of the ten psychiatric and two neurodevelopmental conditions that they worked with. Then, for each psychiatric (not neurodevelopmental) condition the participants had reported they worked with, we asked about the frequency with which patients/clients and their relatives ask about genetic and environmental risk for their condition and their opinion on the value of psychiatric genetic counselling for patients/clients. Further information on these survey items are in the Supplementary Material.

### Overall sample

We describe the overall sample’s genetic knowledge, their perception of genetic and environmental risk for each psychiatric condition, how often patients/clients ask about genetic or environmental risk for each psychiatric condition, and their support for a psychiatric genetic counselling service. We used a paired samples t-test to compare confidence in discussing genetic versus environmental risk.

### Comparing mental health professionals with vs. without a medical degree Training background of participants

Based on their current role, we categorised the participants as having a medical degree or not. Roles included in ‘medical degree’ were psychiatrists and General Practitioners (GPs). Roles included in ‘no medical degree’ were clinical psychologists, assistant psychologists, associate psychologists, children’s wellbeing practitioners, clinical wellbeing psychologists, counselling psychologists, counsellors, educational mental health practitioners, educational psychologists, mental health nurses, and physical wellbeing leads.

### Statistical tests

First, we conducted unadjusted linear regressions to explore whether having a medical degree (versus not) was associated with 1) genetic knowledge (measured via 6-item iGLAS) 2) how confident participants feel discussing a) genetic risk and b) environmental risk with their patients/clients or their relatives. Next, given that Chapman et al.^13^ found an unexpected significant sex difference in genetic knowledge and that 50% of those with a medical degree were male versus only 20% of those without a medical degree in our sample, we performed adjusted models where we controlled for participants’ assigned-sex-at-birth. We accounted for multiple testing with a Bonferroni-corrected *p-*value threshold adjusted for the overall number of statistical tests 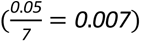.

### Ethical approval

The PerPsych study was approved by the Psychiatry, Nursing, and Midwifery Research Ethics Subcommittee at King’s College London on 24th June 2021 (HR/DP-20/21-22019). Written informed consent was obtained from all participants.

## Results

### Sample

We received survey responses from 152 mental health professionals, both qualified and in-training, in a range of roles and professional settings (**Table 1; Supplementary Table 1**). Overall, the sample had a mean age of 34 years (SD=10). The majority reported being white (82%), identified as a woman (70%), and only worked for the NHS (73%). Those who were not in training qualified between 1983 and 2022. A greater proportion of participants without a medical degree identified as a woman (80%) than those with a medical degree (50%).

**Table 1.**
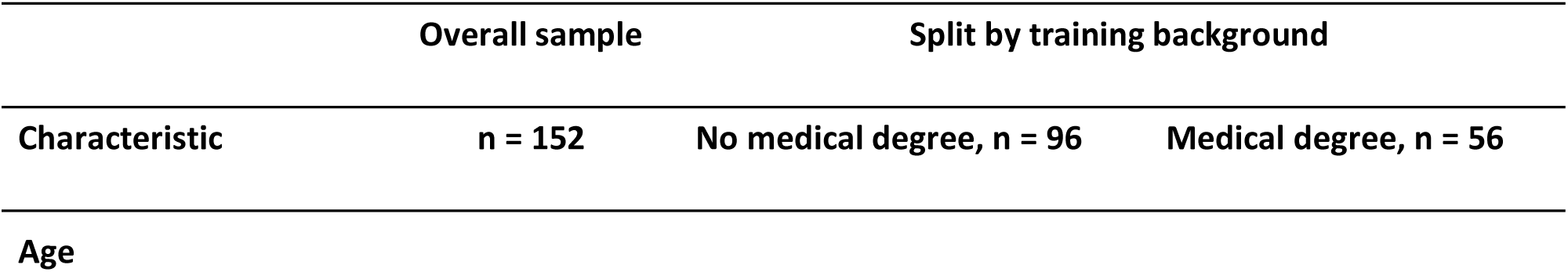
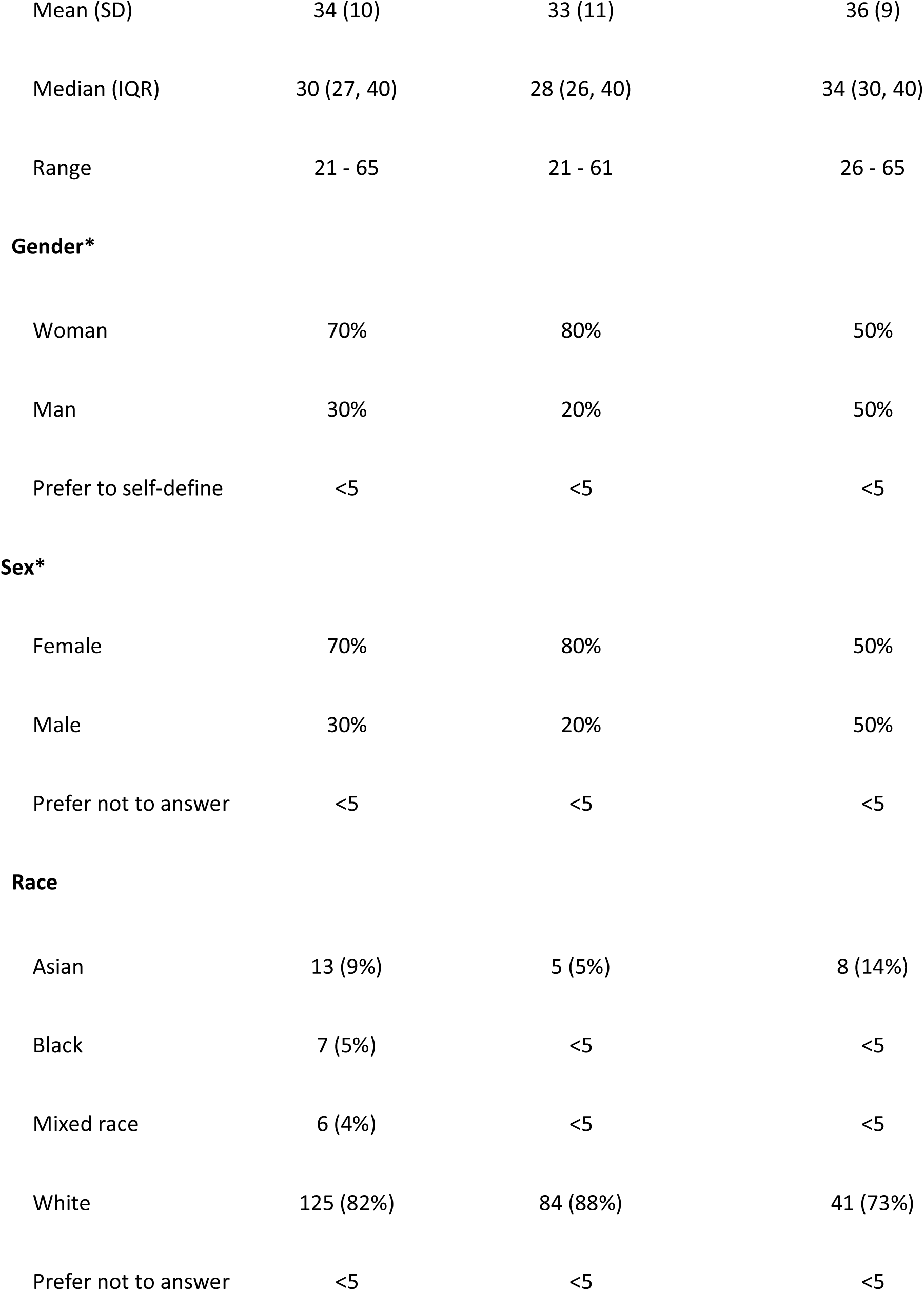
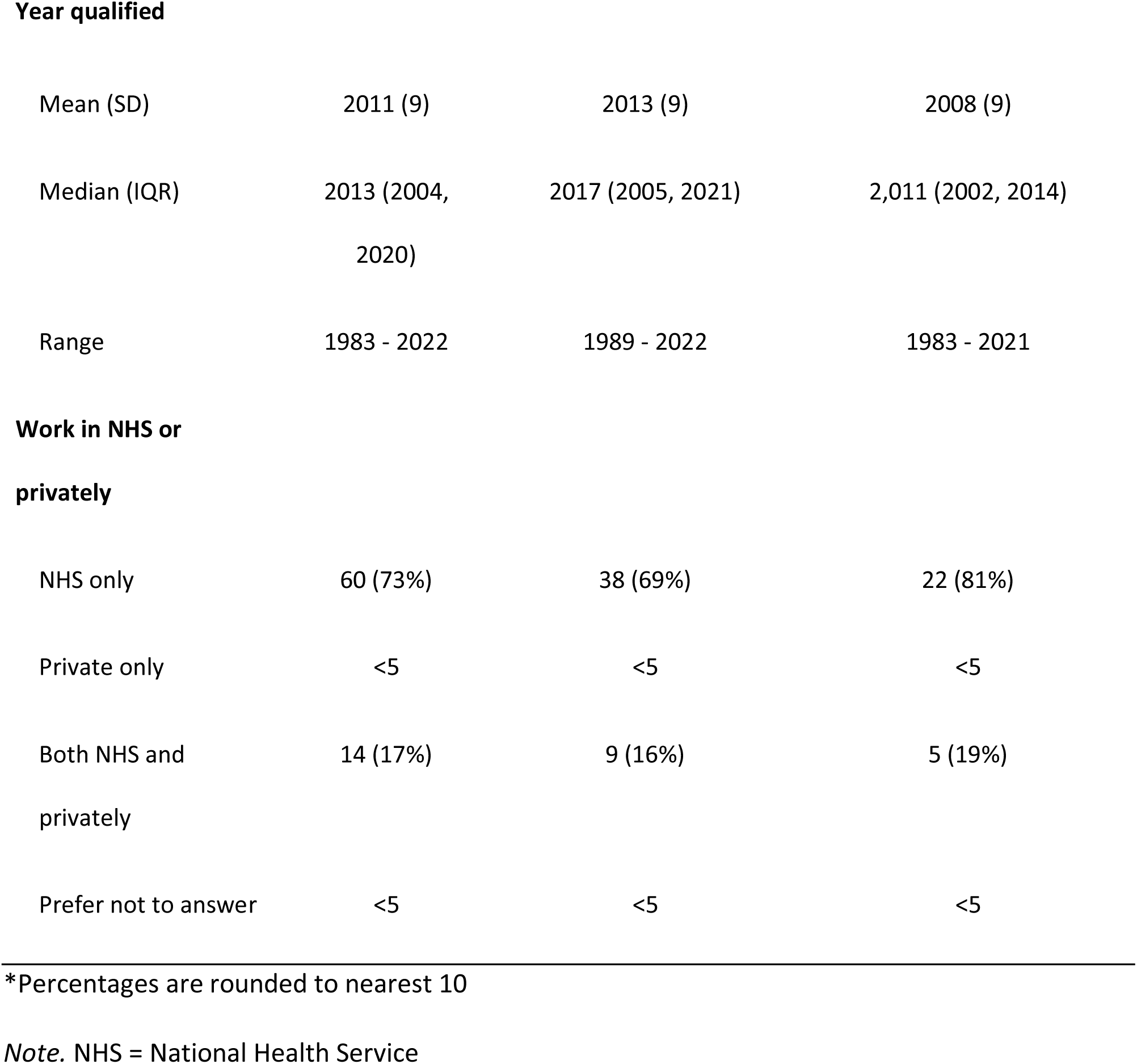
Sample descriptive statistics (total n = 152)

|  | Overall sample |  | Split by training background |
| --- | --- | --- | --- |
| Characteristic | n = 152 | No medical degree, n = 96 | Medical degree, n = 56 |
| Age |  |  |  |
| Mean (SD) | 34 (10) | 33 (11) | 36 (9) |
| Median (IQR) | 30 (27, 40) | 28 (26, 40) | 34 (30, 40) |
| Range | 21 - 65 | 21 - 61 | 26 - 65 |
| **Gender\*** |  |  |  |
| Woman | 70% | 80% | 50% |
| Man | 30% | 20% | 50% |
| Prefer to self-define | <5 | <5 | <5 |
| **Sex\*** |  |  |  |
| Female | 70% | 80% | 50% |
| Male | 30% | 20% | 50% |
| Prefer not to answer | <5 | <5 | <5 |
| **Race** |  |  |  |
| Asian | 13 (9%) | 5 (5%) | 8 (14%) |
| Black | 7 (5%) | <5 | <5 |
| Mixed race | 6 (4%) | <5 | <5 |
| White | 125 (82%) | 84 (88%) | 41 (73%) |
| Prefer not to answer | <5 | <5 | <5 |
| **Year qualified** |  |  |  |
| Mean (SD) | 2011 (9) | 2013 (9) | 2008 (9) |
| Median (IQR) | 2013 (2004,<br>2020) | 2017 (2005, 2021) | 2,011 (2002, 2014) |
| Range | 1983 - 2022 | 1989 - 2022 | 1983 - 2021 |
| **Work in NHS or privately** |  |  |  |
| NHS only | 60 (73%) | 38 (69%) | 22 (81%) |
| Private only | <5 | <5 | <5 |
| Both NHS and<br>privately | 14 (17%) | 9 (16%) | 5 (19%) |
| Prefer not to answer | <5 | <5 | <5 |

\*Percentages are rounded to nearest 10
*Note.* NHS = National Health Service

#### Descriptives

##### Knowledge of genetics

A total of 137 participants answered the 6-item iGLAS. On average, participants scored 4.36 out of the maximum 6 points (SD=1.28), i.e., 73% correct. This is comparable with the average percentage correct answers of 66% found in 5,310 participants who completed the 18-item pilot version of the iGLAS^13^. **Supplementary Table 2** compares the percentage correct across five questions in our sample to participants from Chapman et al., (2019) separated by profession into undergraduate psychology students (n=112), legal practitioners (n=90), and teachers (n=244). As expected, out of all professionals in **Supplementary Table 2**, those with a medical degree scored highest on all questions bar one in which they scored similarly (63% correct) to legal practitioners (70%) and teachers (66%), whilst those without a medical degree scored most similarly to teachers. Importantly, 100% of participants with a medical degree and 86% of those without gave the correct answer to the question ‘Genetic contribution to the risk for developing Schizophrenia comes from: a) One gene, b) Many genes’, far higher than that of legal practitioners (52%), teachers (62%), and students (58%). Twenty-three percent of the sample answered all six questions correctly, no participants answered all incorrectly, and 92% percent answered at least half correctly. Notably, 12 individuals (9%) answered that the genetic contribution to the risk for schizophrenia comes from a single gene (all of whom were those without a medical degree).

### Training background comparison

Participants with a medical degree scored significantly higher (mean=5.02, SD=0.91) than those without a medical degree (mean=3.97, SD=1.31, β=1.05, 95% CI=0.64, 1.47, *p*=<0.001), and this remained significant after controlling for the participants’ sex (β=0.96, 95% CI=0.52, 1.39, *p*=<0.001).

#### Perception of genetic and environmental contributions to twelve psychiatric conditions

For each condition, participants gave a range of answers; however there was large variation in the frequency of each answer across the conditions. Notably, 15 individuals (10%) said that at least one of the psychiatric conditions was caused by ‘only environmental factors’ or ‘only genetic factors’. Some participants perceived PTSD (n<5) and personality disorders (n<5) to be caused by ‘only environmental factors’, while some perceived autism (n=8, 8%), ADHD (n<5) and anxiety (n<5) to be caused by ‘only genetic factors’ (**Figure 1**). The mean ratings of confidence in their answers (score of 0 to 100%) across the twelve conditions ranged from 67% (SD=23%) for ADHD to 79% (SD=14%) for PTSD (**Supplementary Table 3**).

**Figure 1.**
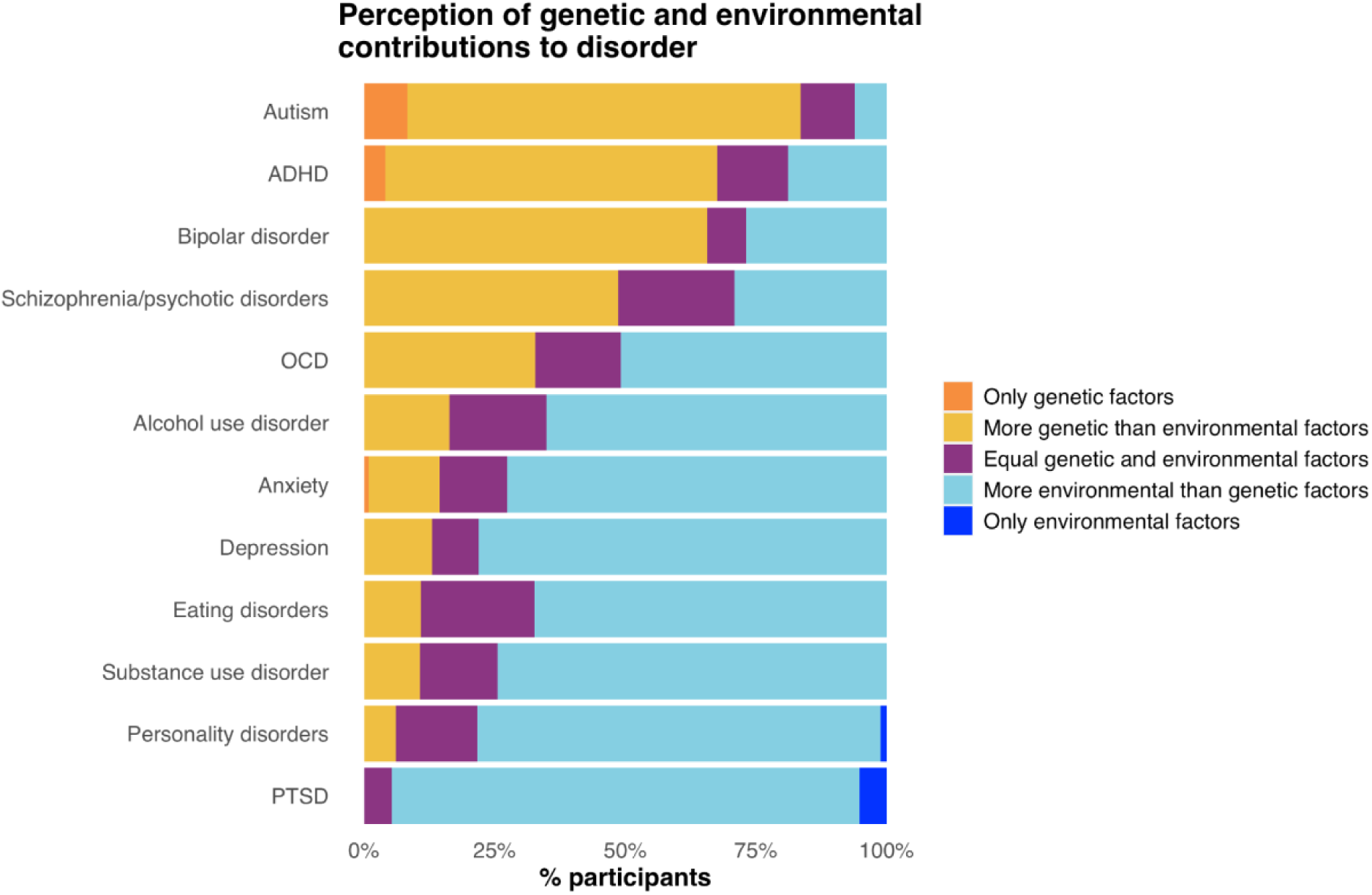
<u>Mental health professionals’ perception of genetic and environmental contributions to psychiatric conditions</u> Question: Please select an option according to how much you think [condition] is caused by inherited genetic factors and/or environmental factors. Answer options: ‘only environmental factors’, ‘almost only environmental factors’, ‘mainly environmental factors’, ‘slightly more environmental factors’, ‘equal genetic and environmental factors’, ‘slightly more genetic factors’, ‘mainly genetic factors’, ‘almost only genetic factors’, ‘only genetic factors’. For the purpose of this plot, ‘almost only genetic factors’, ‘mainly genetic factors’, ‘slightly more genetic factors’ were grouped together as ‘more genetic than environmental factors’ and ‘almost only environmental factors’, ‘mainly environmental factors’, ‘slightly more environmental factors’ were grouped together as ‘more environmental than genetic factors’. The conditions are ordered based on the descending frequency of the proportion who answered ‘only genetic factors’ and ‘more genetic than environmental factors’.

### Training background comparison

A higher proportion of participants without a medical degree selected ‘only genetic factors’ or ‘only environmental factors’ when asked about the cause(s) of the ten psychiatric conditions. Out of 94 participants without a medical degree, 13 (14.1%) selected these options, compared to <5 out of 56 (<9%) participants with a medical degree. Due to low cell counts, we were not able to test whether this difference was statistically significant.

### Patients/clients: Frequency with which they ask about genetic and environmental risk

#### Questions about genetic predisposition

The proportion of participants who reported that their patients/clients ‘always’ or ‘often’ ask about their genetic predisposition ranged from <6% (in those with PTSD, n<5) to 32% (in those with bipolar disorder). Twenty-seven percent of the sample indicated that their patients/clients ‘always’ or ‘often’ asked about their genetic predisposition for at least one condition (**Figure 2**). The proportion who reported that their patients/clients ‘never’ or ‘rarely’ asked ranged from 32% in bipolar disorder to 92% in PTSD. Seventy-one percent said that their patients/clients ‘never’ or ‘rarely’ asked about their genetic predisposition for at least one condition.

**Figure 2.**
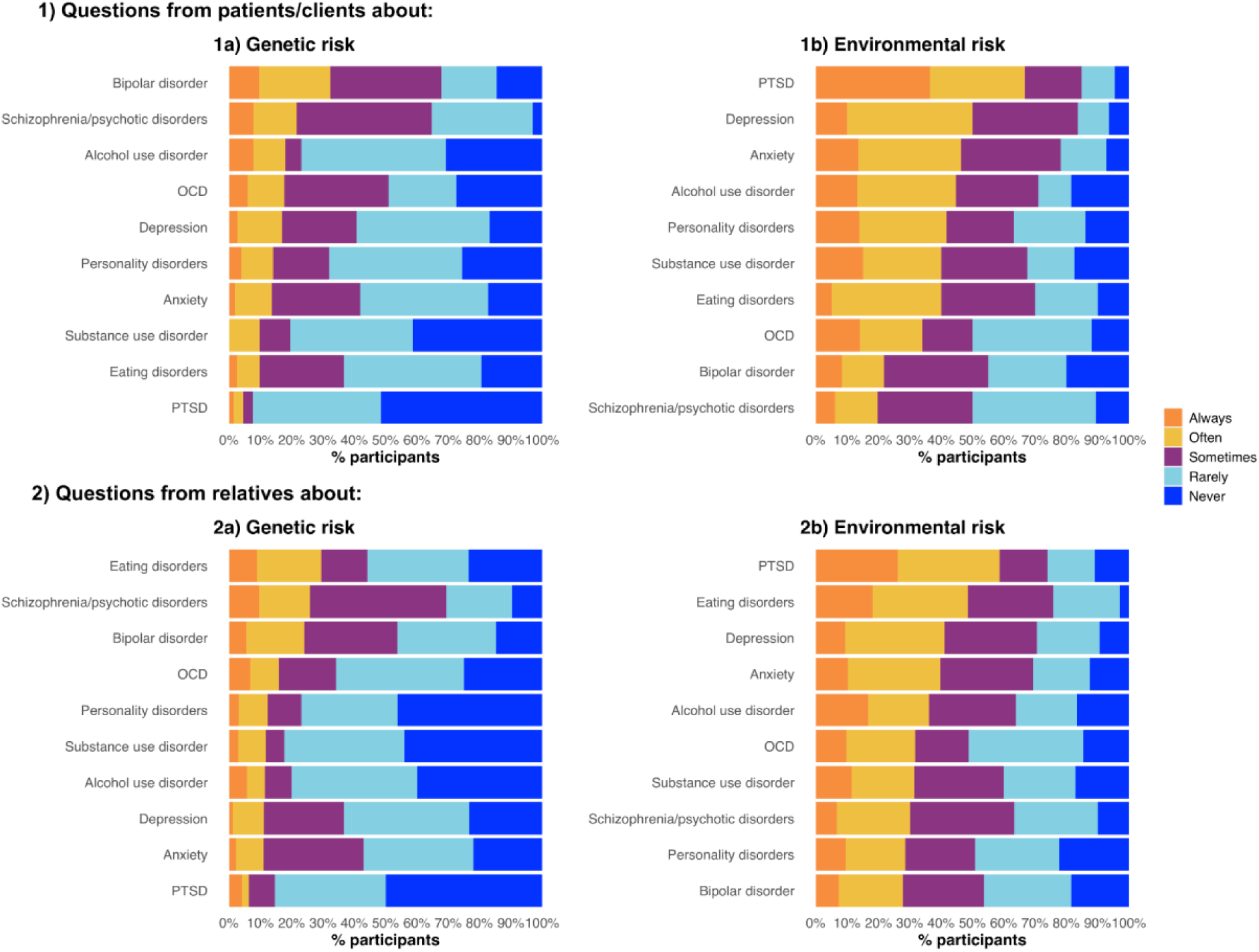
Frequency with which mental health professionals are asked about genetic predisposition and environmental risk for psychiatric conditions by patients/clients (1a and 1b) and their relatives (2a and 2b) *Question (1a and 1b): Please indicate how often your patients/clients (with the disorders below) raise questions about their [genetic predisposition/environmental risk] for that psychiatric disorder. Question (2a and 2b): Please indicate how often the relatives of your patients/clients (with the disorders below) raise questions about their [genetic predisposition/environmental risk] for that psychiatric disorder. The conditions are ordered based on the descending frequency of the proportion who answered ‘always’ and ‘often’*.

#### Questions about environmental risk

In general, mental health professionals reported being more commonly ‘always’ or ‘often’ asked about the environmental contributions to each psychiatric condition compared to their genetic predisposition. The proportion of participants who said that their patients/clients ‘always’ or ‘often’ asked questions ranged from 20% in patients/clients with schizophrenia to 67% in patients/clients with PTSD. Fifty-six percent indicated that their patients/clients ‘always’ or ‘often’ asked about the environmental contributions for at least one condition. The proportion who said that patients/clients ‘never’ or ‘rarely’ asked ranged from 15% in patients/clients with PTSD to 50% in patients/clients with schizophrenia (**Figure 2**). Forty-five percent said that their patients/clients ‘never’ or ‘rarely’ asked about environmental risk for at least one condition.

### Relatives: Frequency with which they ask about genetic and environmental risk

#### Questions about genetic predisposition

The proportion of participants who reported that the *relatives* of patients/clients ‘always’ or ‘often’ asked about the genetic predisposition ranged from <5 in relatives of individuals with PTSD to 29% in relatives of individuals with eating disorders. The proportion who reported that relatives ‘never’ or ‘rarely’ asked about genetics ranged from 31% in relatives of individuals with schizophrenia to 85% in relatives of individuals with PTSD.

#### Questions about environmental risk

The proportion of participants who reported that the relatives of patients/clients ‘always’ or ‘often’ asked about environmental risk ranged from 28% in relatives of individuals with bipolar disorder to 59% in relatives of individuals with PTSD. The proportion who reported that relatives ‘never’ or ‘rarely’ asked about environmental risk ranged from 24% in relatives of individuals with eating disorders to 51% in relatives of individuals with OCD.

#### Confidence discussing genetic and environmental risk with patients/clients or relatives

Overall, mental health professionals reported feeling more confident discussing environmental risk with patients/clients or their relatives than they feel discussing genetic risk; only 30% agreed or strongly agreed that they were confident discussing genetic risk compared to 80% for environmental risk (**Figure 3**). A paired samples Wilcoxon signed rank test in which the answers were assigned numeric values on a 5-point scale (i.e., ‘strongly disagree’=0 to ‘strongly agree’=4) showed that participants had significantly greater confidence discussing environmental risk (median=3, IQR=1) than genetic risk (median=1, IQR=2), V=4522, *p<* 0.001.

**Figure 3.**
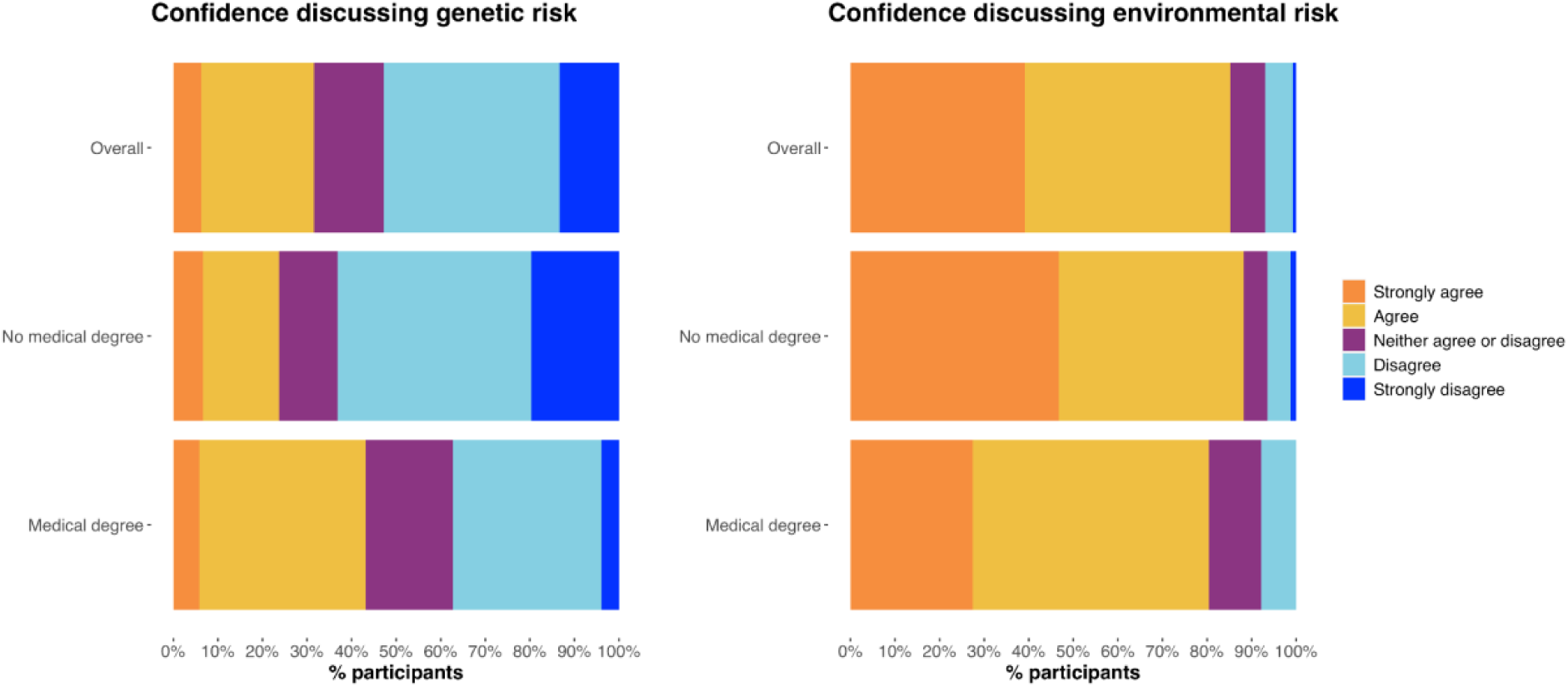
Mental health professionals’ agreement/disagreement with whether they feel confident discussing genetic (left) and environmental (right) risk for psychiatric conditions with patients/clients or their relatives *Question: To what extent do you agree with this statement: I feel confident discussing environmental risk with my patients/clients or the relatives of my patients/clients*.

### Training background comparison

A total of 43% with a medical degree agreed or strongly agreed that they feel confident discussing genetic risk compared to 24% without a medical degree (**Figure 3**). After assigning the answers numeric values on a 5-point scale as above, those with a medical degree feel statistically significantly more confident discussing genetic risk than those without a medical degree (β=0.60, 95% CI=0.20, 1.01, *p*=0.004) however this was non-significant after controlling for the participants’ sex (β=0.37, 95% CI=-0.04, 0.78, *p*=0.08). A total of 80% with a medical degree agreed or strongly agreed that they feel confident discussing environmental risk compared to 89% without a medical degree (**Figure 3**). After assigning the answers numeric values as above, we found that having a medical degree was not associated with confidence discussing environmental risk (β=-0.27, 95% CI=-0.58, 0.04, *p*=0.08), including after controlling for sex (β=-0.38, 95% CI=-0.70, -0.05, *p*=0.02).

### Support for a psychiatric genetic counselling service

We found generally high agreement that a psychiatric genetic counselling service would be helpful across all ten psychiatric conditions included in the survey. Mental health professionals thought it would be most helpful for individuals with eating disorders (81% agreed), closely followed by individuals with schizophrenia/psychotic disorders (80% agreed), and then bipolar disorder (76% agreed). Participants thought it would be the least helpful for individuals with PTSD (50% agreed) (**Supplementary** Figure 1).

Similarly, there was high agreement that the mental health professionals would refer their patients/clients for psychiatric genetic counselling (**Supplementary** Figure 2).

## Discussion

We surveyed 152 trainee and qualified mental health professionals in the UK about their genetic knowledge, perceptions of genetic and environmental risk for psychiatric conditions, the frequency that they are asked questions about these factors, and how confident they feel discussing these with patients/clients and their relatives.

### Knowledge of genetics

Genetic knowledge was generally good (the mean number of correct answers was 4.36 out of 6, or 73%), with only a small proportion of the sample (8%) answering more than half of the questions incorrectly. Encouragingly, no participants answered all of the questions incorrectly. Participants without a medical degree displayed a significantly lower understanding of genetics compared to participants with a medical degree. In the UK, mental health educational programmes such as clinical psychology doctorates or mental health nursing undergraduate degrees rarely include substantive teaching on psychiatric genetics. Nonetheless, we do not suggest extensively re-designing such training programmes; many existing tools can act as a useful starting point^30,36^. Indeed, the ISPG recently called for inclusive psychiatric genetics education for all mental health clinicians, starting with the utilisation of existing free online resources such as those of the National Human Genome Research Institute^37^. Our paper supports and extends the ISPG’s call to action by demonstrating its support from mental health professionals (as shown by the high agreement from participants that discussion of genetic risk would be beneficial for patients/clients) and its relevance in the UK.

### Perception of genetic and environmental contributions to twelve psychiatric conditions

We found variation in the perceived causes of twelve psychiatric and neurodevelopmental conditions, with two notable findings. First, the neurodevelopmental conditions ADHD and autism, were most often perceived as being more genetically than environmentally caused, consistent with research suggesting that reluctance to disclose an autism diagnosis relates to its perceived link to identity^38^. Second, all other conditions were more often perceived as caused by more genetic than environmental factors in comparison to PTSD, despite some of them (e.g., depression and anxiety) having comparable, or in some cases lower, heritability estimates^39^. For PTSD, the requirement of trauma exposure for diagnosis may anchor perceptions toward environmental explanations, illustrating how diagnostic criteria can shape perceptions of causes.

Whilst the majority of the mental health professionals demonstrated an understanding that psychiatric and neurodevelopmental conditions are caused by a combination of genetic and environmental factors, 10% indicated that they believed at least one condition they worked with is caused by only environmental factors or only genetic factors. Descriptively, those without a medical degree were more likely to hold such absolute views than those with a medical degree. Further, 9% of participants, all without a medical degree, endorsed a belief that the genetic contribution to schizophrenia originates from a single gene. Identifying that genetically (and environmentally) absolute beliefs towards psychiatric conditions are held even by a small minority of those specially trained to treat them highlights key gaps in mental health professional education systems. As the field of psychiatric genetics advances, it is crucial that we provide new educational opportunities, especially for those trained in disciplines other than medicine.

### Frequency with which patients/clients and their relatives ask about genetic and environmental risk

While our findings show that mental health professionals are more commonly asked about environmental risk factors compared to genetic risk factors (aside from in bipolar disorder), they confirm that those affected by mental illness and their families are also interested in genetic risk. Indeed, we found that a sizable but varying proportion of participants are *often* or *always* asked about condition-specific genetic risk, emphasising the importance of adequate training in the genetic underpinnings of psychiatric conditions, particularly bipolar disorder, schizophrenia/psychotic disorders, and eating disorders. We found high agreement that psychiatric genetic counselling could be helpful for a number of the disorders, especially eating disorders, schizophrenia/psychotic disorders, and bipolar disorder. This finding strengthens the argument that patients and their families have an interest in understanding the role of genetic factors in shaping individual susceptibility to psychiatric disorders. In the UK, there is just one psychiatric genomics service - the All Wales Psychiatric Genomics Service. While genetic healthcare practitioners have acknowledged its potential for benefitting patients, several implementation barriers have also been noted, including the need for specific training and the potential to cause negative emotions^40^. Therefore, our findings should be interpreted in the context of psychiatric genetic counselling in the UK, which remains in a nascent stage and requires further research.

### Confidence discussing genetic and environmental risk with patients/clients or relatives

Overall the respondents reported significantly lower confidence in discussing genetic risk than that of environmental risk; 80% agreed that they feel confident discussing environmental risk, while only 30% agreed for genetic risk. This aligns with previous research that found that 23% of 844 psychiatrists in the USA and Canada feel ‘competent’ to discuss genetics with patients, and an even smaller proportion (15%) feel ‘sufficiently equipped’ to do so^41^. Whilst such a finding could be attributed to low confidence discussing *general* risk factors for psychiatric conditions, our additional assessment of environmental risk, which the vast majority of participants reported being confident discussing, indicates that this is specific to genetics and thus likely related to a distinct training gap. We add nuance to these previous studies by demonstrating that the training gap is more evident in education outside of a medical degree; those with a medical degree were significantly more confident discussing genetics than those without a medical degree, but we did not observe this for environmental risk.

Additionally, our findings stress a distinction between having a basic understanding that both genes and environment contribute to psychiatric disorders and having the skills and confidence to communicate that knowledge in a therapeutic way. Genetic counsellors discuss the genetic basis of disease with those affected, or those at risk of being affected, within a psycho-therapeutic environment to help them come to terms with the medical, psychological, and familial implications of the genetic contribution to the disease in question^42^. Genetic counselling for psychiatric conditions is effective for both psychological/emotional and knowledge outcomes^43^. Individuals may approach a mental health professional with questions about psychiatric genetic risk for a number of reasons, e.g., seeking clarification about their own risk after a family member has been diagnosed with a psychiatric condition; after receiving the results of a DTC genetic test; they might be concerned that their children could develop the condition they have themselves, among others. All of these individuals deserve to have their questions answered accurately and sensitively. While genetic counsellors are specifically trained to do this, as of 2019, there are only around 310 in the UK^22^ and they do not usually work within psychiatry. Therefore, individuals with a mental illness rarely get the opportunity to speak to a genetic counsellor as part of their treatment. Rather, as our findings show, mental health professionals who are often having these conversations in the clinic do not feel confident doing so. Utilising existing resources to help mental health professionals better understand psychiatric genetics is a vital first step; future research should investigate how to best convey such information to ensure that the conversation is informative and empowering. For example, a recent study found that both patients and primary care practitioners better understood polygenic scores when represented as a continuous rather than binary dimension^44^.

### Strengths and limitations

This study is the first, to our knowledge, to explore how often mental health professionals are asked about psychiatric genetic risk, enabling us to demonstrate that patients/clients and their families are indeed interested in learning more about how genetics may have influenced the development of their, or their family member’s, condition. Whilst our sample size was small, we highlight the difficulty in recruiting mental health clinicians given the particularly high demands on their time. Further, our recruitment approach meant we were limited to those who were active on social media or email. However, we hope that our offer of a potential prize for participation (an iPad) meant that we also recruited a broader sample including those not motivated by a pre-existing interest in psychiatric genetics. Given that we wanted to make the length of our survey as short as possible, some of our questions were brief and therefore likely did not capture our measures in their full complexity and nuance. For example, interpretations of ‘genetics’ may differ and depending on your understanding, a conversation about family history may or may not have been considered relevant. Future research would benefit from the collection of more detailed data, e.g. via interviews with mental health professionals. An additional limitation is that we made an assumption about their training background because we did not explicitly ask whether they had a medical degree or not, which may have meant some participants were misclassified if they had switched careers.

## Conclusions

Our study shows that individuals with psychiatric conditions and their family members are curious about the genetic contribution to their condition(s) and ask the professionals treating them questions about this. Yet, mental health professionals do not feel confident having these conversations. Previous research has shown that understanding the causes of psychiatric conditions can bring positive emotional and behavioural outcomes for individuals living with them and, given that mental health professionals are well placed to have these conversations, it is vital that they are trained to do so properly.

## Data Availability

PerPsych study data are available via a data request application to the NIHR BioResource (https://bioresource.nihr.ac.uk/using-our-bioresource/academic-and-clinical-researchers/apply-for-bioresource-data/). The data are not publicly available due to restrictions outlined in the study protocol and specified to participants during the consent process.

## Acknowledgements

We thank all participants for taking time to complete the survey as part of this study. We gratefully acknowledge Professor J9 Austin for their guidance on this project.

## Ethics statement

The authors assert that all procedures contributing to this work comply with the ethical standards of the relevant national and institutional committees on human experimentation and with the Helsinki Declaration of 1975, as revised in 2013. The PerPsych study was approved by the Psychiatry, Nursing, and Midwifery Research Ethics Subcommittee at King’s College London on 24th June 2021 (HR/DP-20/21-22019).

## Author contribution

*Data curation*: Helena L. Davies, Jessica Mundy, Molly R. Davies. *Data cleaning*: Helena L. Davies, Jessica Mundy, Karla Mohoric. *Formal analysis*: Helena L. Davies, Jessica Mundy, Gerome Breen, Evangelos Vassos. *Funding acquisition*: Thalia C. Eley, Gerome Breen, Evangelos Vassos. *Investigation*: Helena. L. Davies, Jessica Mundy, Gerome Breen, Evangelos Vassos. *Methodology*: Helena L. Davies, Jessica Mundy, Gerome Breen, Evangelos Vassos. *Project administration*: Helena L. Davies, Jessica Mundy, Molly R. Davies. *Resources*: Molly R. Davies, Thalia C. Eley, Gerome Breen. *Supervision*: Gerome Breen, Evangelos Vassos. *Visualization*: Helena L. Davies, Jessica Mundy. *Writing—original draft*: Helena L. Davies, Jessica Mundy. *Writing—review & editing*: Helena L. Davies, Jessica Mundy, Karla Mohoric, Molly R. Davies, Thalia C. Eley, Gerome Breen, Evangelos Vassos. *Preparation*: Helena L. Davies, Jessica Mundy, Gerome Breen, Evangelos Vassos.

