## Supplementary Materials for "UK-based mental health professionals’ perceptions of genetic and environmental risk for psychiatric conditions"

#### Supplementary methods

***Survey items***

We assessed participants' knowledge of genetics with the shortened version of the International Genetic Literacy and Attitudes Survey (43 [6-item version obtained via personal communication]). Additionally, we asked the participants how confident they felt discussing genetic and environmental risk with patients/clients and their relatives by asking them to indicate the extent to which they agreed or disagreed with a number of statements on a 5-point likert scale ('strongly disagree' to 'strongly agree'), for example: 'I feel confident discussing genetic risk with my patients/clients'.

Next, we asked participants their perception of the genetic and environmental contributions to each of the psychiatric and neurodevelopmental conditions that they worked with. Participants responded via a scale ranging from 'only genetic factors' to 'only environmental factors'. Immediately after each question, we asked them to indicate how confident they were in their answer (ranging 0-100%).

Then, for each psychiatric (not neurodevelopmental) condition the participants had reported they worked with, we asked the following questions:

- **Frequency with which patients/clients and their relatives ask about genetic or environmental risk for their condition.** We explored how often participants are asked by patients/clients and their relatives about (1) their genetic predisposition for the psychiatric condition (e.g., 'Please indicate how often your patients/clients with [*psychiatric condition*] raise questions about their genetic predisposition for that psychiatric disorder') and (2) their environmental risk (e.g., 'Please indicate how often your patients/clients with [*psychiatric condition*] raise questions about their environmental risk for that psychiatric disorder').

- **Opinion on value of psychiatric genetic counselling for patients/clients.** Finally, we gave participants a definition of psychiatric genetic counselling - developed with a genetic counsellor (Supplementary Material) - and asked participants to indicate to what extent they agreed with a number of statements on a 7-point Likert scale: 'A psychiatric genetic counselling service would be helpful for my patients/clients with [*psychiatric condition*]' and 'If a psychiatric genetic counselling service was available, I would recommend it to/refer my patients/clients with [*psychiatric disorder*]'. Results are in Supplementary Material.

***Training background of participants***

Roles included in 'medical degree' were psychiatrists and General Practitioners (GPs). Roles included in 'no medical degree' were clinical psychologists, assistant psychologists, associate psychologists, children's wellbeing practitioners, clinical wellbeing psychologists, counselling psychologists, counsellors, educational mental health practitioners, educational psychologists, mental health nurses, and physical wellbeing leads.

***Statistical tests comparing training background***

For the purpose of statistical tests comparing professionals with and without a medical degree, we re-coded the categorical answers to 'I feel confident discussing [genetic/environmental risk] with my patients/clients' into numeric variables: 'strongly disagree' = 0, 'disagree' = 1, 'neither agree or disagree' = 2, 'agree' = 3, 'strongly agree' = 4 so that higher scores represented higher levels of confidence.

### Supplementary tables

Supplementary Table 1. Sample characteristics about the disorders that the mental health professionals work with, their role, and their professional setting (n = 152)

| | **Overall sample** | **Split by training background** | |
|---|---|---|---|
| | **n = 152** | **No medical degree, n = 96** | **Medical degree, n = 56** |
| **Psychiatric conditions worked with*** | | | |
| Depressive disorders | 123 (81%) | 74 (77%) | 49 (88%) |
| Anxiety disorders | 126 (83%) | 81 (84%) | 45 (80%) |
| Posttraumatic stress disorder | 77 (51%) | 51 (53%) | 26 (46%) |
| Bipolar disorder | 67 (44%) | 28 (29%) | 39 (70%) |
| Schizophrenia/psychotic disorders | 74 (49%) | 29 (30%) | 45 (80%) |
| Obsessive compulsive disorder | 57 (38%) | 35 (36%) | 22 (39%) |
| Eating disorders | 47 (31%) | 27 (28%) | 20 (36%) |
| Substance use disorders | 47 (31%) | 24 (25%) | 23 (41%) |
| Personality disorders | 85 (56%) | 39 (41%) | 46 (82%) |
| Alcohol use disorders | 43 (28%) | 20 (21%) | 23 (41%) |
| Attention deficit hyperactivity disorder | 76 (50%) | 48 (50%) | 28 (50%) |
| Autism spectrum disorders | 98 (64%) | 65 (68%) | 33 (59%) |
| Other | 7 (5%) | <5 | <5 |
| **Career field** | | | |
| Assistant or associate psychologist | 20 (14%) | 20 (23%) | 0 (0%) |
| Clinical psychologist | 35 (24%) | 35 (40%) | 0 (0%) |
| Counsellor or therapist | 15 (10%) | 15 (17%) | 0 (0%) |
| Educational psychologist | 5 (3%) | 5 (6%) | 0 (0%) |

| | | | |
|---|---|---|---|
| Mental health nurse | 5 (3%) | 5 (6%) | 0 (0%) |
| Psychological Wellbeing Practitioner or Children's Wellbeing Practitioner | 7 (5%) | 7 (8%) | 0 (0%) |
| General Practitioner (GP) | 6 (4%) | 0 (0%) | 6 (11%) |
| Psychiatrist | 50 (35%) | 0 (0%) | 50 (89%) |
| **Professional setting*** | | | |
| Child and adolescent | 34 (26%) | 27 (36%) | 7 (13%) |
| Adult | 75 (58%) | 41 (54%) | 34 (64%) |
| Old age | 11 (9%) | <5 | NA** |
| General/all ages | <5 | 0 (0%) | <5 |
| Not specialised yet (in-training) | 6 (5%) | <5 | <5 |

*Note: 'Other' in 'Career field' includes social workers, education mental health practitioners, and physical wellbeing leads.*
**participants could select more than one answer*
***value suppressed*

Supplementary Table 2. Comparison of responses to 6-item iGLAS from Chapman et al. (2019) to our sample (n = 152)

| | Total sample (n = 5310) | Legal Practitioner (n = 90) | Teachers (n = 244) | Undergrad Psychology students (n = 112) | Our total sample (n = 152) | No medical degree (n = 96) | Medical degree (n = 56) |
|---|---|---|---|---|---|---|---|
| **What is a genome?**<br><br>o A sex chromosome<br>*o The entire sequence of an individual's DNA*<br>o All the genes in DNA<br>o Gene expression | 53% | 54% | 61% | 55% | 72% | 64% | 84% |
| **Which of the following 4 letter groups represent the base units of DNA?**<br><br>o GHPO<br>o HTPR<br>*o GCTA*<br>o LFWE | 76% | 54% | 67% | 66% | 80% | 69% | 98% |
| **On average, how much of their total DNA is the same in two people selected at random?**<br><br>o Less than 50%<br>o 75%<br>o 90%<br>*o More than 99%* | 60% | 47% | 50% | 35% | 54% | 48% | 64% |
| **The DNA sequence in two different cells, for example a neuron and a heart cell, of one person, is:**<br><br>o Entirely different<br>o About 50% the same<br>o More than 90% the same<br>*o 100% identical* | 74% | 70% | 66% | 38% | 50% | 43% | 63% |

| | | | | | | | |
|---|---|---|---|---|---|---|---|
| **Genetic contribution to the risk for developing Schizophrenia comes from:** | 67% | 52% | 62% | 58% | 91% | 86% | 100% |
| o One gene | | | | | | | |
| *o Many genes* | | | | | | | |
| **Some of the genes that relate to dyslexia also relate to Attention Deficit Hyperactivity Disorder (ADHD):** | NA | NA | NA | NA | | | |
| *o True* | | | | | | | |
| o False | | | | | 89% | 88% | 92% |

Supplementary Table 3. Mental health professionals' confidence in their perception of the genetic and environmental causes of psychiatric conditions

| | **n = 152** |
|---|---|
| **Depression** | |
| Mean (SD) | 70 (16) |
| Median (IQR) | 70 (64, 80) |
| Range | 0 - 100 |
| **Anxiety** | |
| Mean (SD) | 69 (16) |
| Median (IQR) | 70 (60, 80) |
| Range | 16 - 100 |
| **PTSD** | |
| Mean (SD) | 79 (14) |
| Median (IQR) | 80 (70, 90) |
| Range | 40 - 100 |
| **Bipolar disorder** | |
| Mean (SD) | 71 (16) |

| | |
|---|---|
| Median (IQR) | 72 (60, 80) |
| Range | 29 - 100 |
| **Schizophrenia/psychotic disorders** | |
| Mean (SD) | 71 (17) |
| Median (IQR) | 70 (62, 82) |
| Range | 15 - 100 |
| **OCD** | |
| Mean (SD) | 69 (17) |
| Median (IQR) | 70 (59, 80) |
| Range | 29 - 100 |
| **Eating disorders** | |
| Mean (SD) | 71 (17) |
| Median (IQR) | 70 (61, 85) |
| Range | 26 - 100 |
| **Personality disorders** | |
| Mean (SD) | 71 (19) |
| Median (IQR) | 73 (60, 83) |
| Range | 10 - 100 |
| **Substance use disorder** | |
| Mean (SD) | 71 (19) |
| Median (IQR) | 75 (60, 84) |
| Range | 15 - 100 |
| **Autism** | |
| Mean (SD) | 68 (22) |
| Median (IQR) | 71 (56, 82) |
| Range | 5 - 100 |

| | |
|---|---|
| **Alcohol use disorder** | |
| Mean (SD) | 72 (19) |
| Median (IQR) | 75 (63, 84) |
| Range | 28 - 100 |
| **ADHD** | |
| Mean (SD) | 67 (23) |
| Median (IQR) | 71 (56, 81) |
| Range | 0 - 100 |

### Supplementary figures

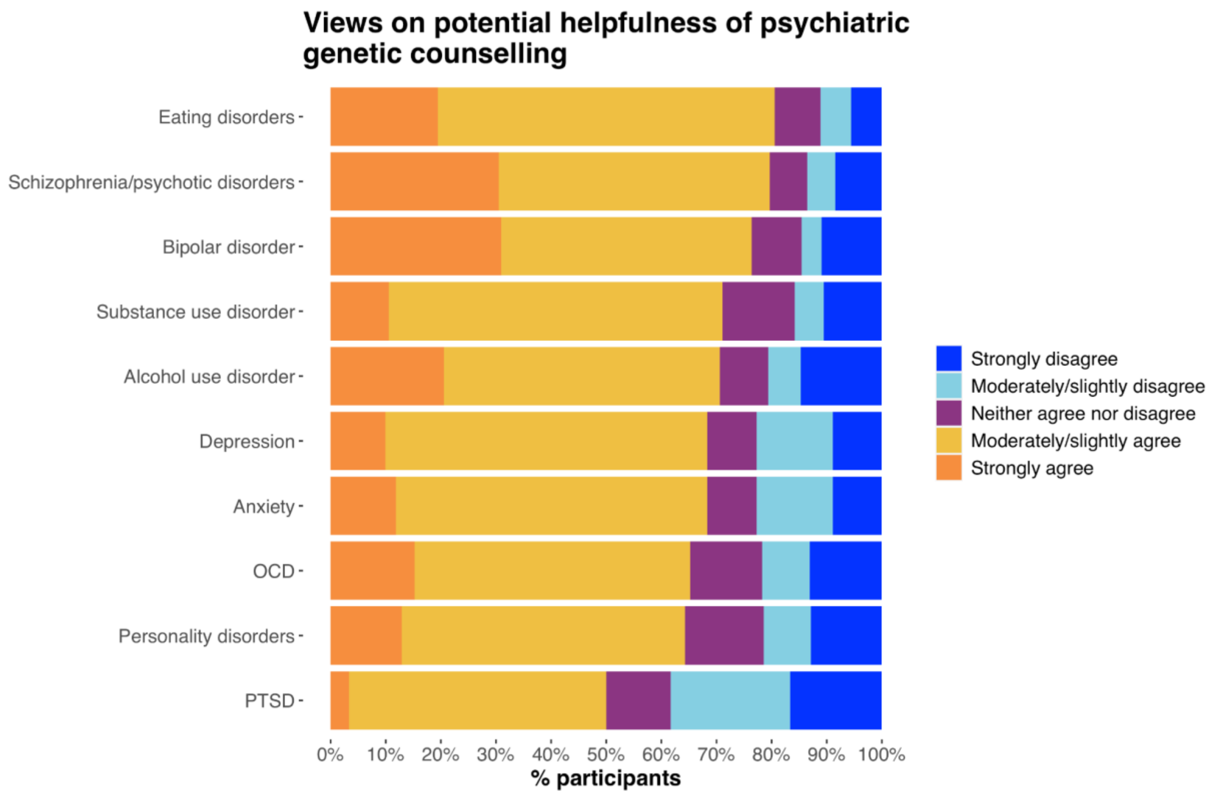


<u>Supplementary Figure 1. Mental health professionals' agreement/disagreement about whether psychiatric genetic counselling would be helpful for their patients/clients with a certain psychiatric disorder.</u>

*Question: "To what extent do you agree with this statement: A psychiatric genetic counselling service would be helpful for my patients/clients with [condition]. Conditions are ordered by proportion who answered with "'strongly agree' or 'moderately/slightly agree'.*

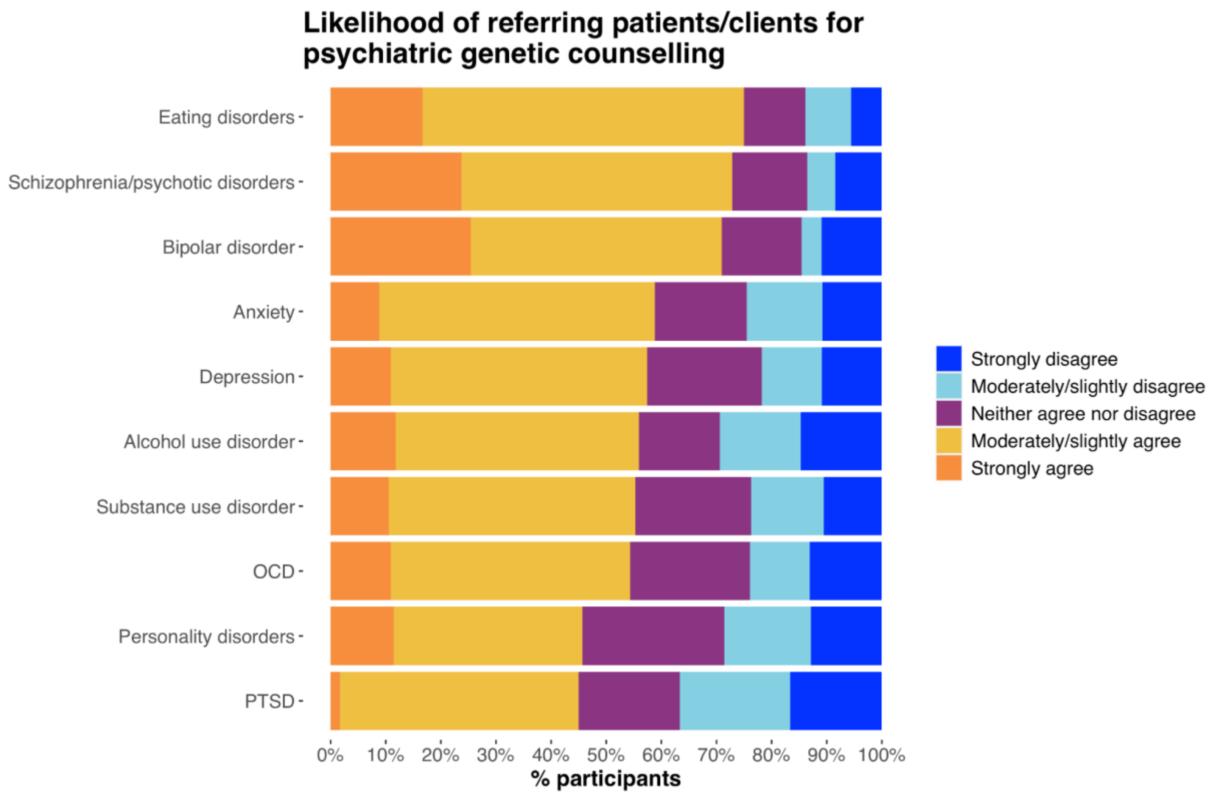


Supplementary Figure 2. Mental health professionals' agreement/disagreement about whether they would refer their patients/clients to a psychiatric genetic counselling service.

*Question: To what extent do you agree with this statement: I would refer my patients/clients with [condition] to a psychiatric genetic counselling service. Conditions are ordered by proportion who answered with 'strongly agree' or 'moderately/slightly agree'.*
